# Recommendations on surveillance for recurrence in asymptomatic patients following surgery in 16 common cancer types: a systematic review

**DOI:** 10.1101/2024.09.19.24313975

**Authors:** Hannah Harrison, Bhumi K. Shah, Faris Khan, Carley Batley, Chiara Re, Sabrina H. Rossi, Georgia Stimpson, Eamonn Gilmore, Ellie White, Sofia Kler-Sangha, Aufia Espressivo, Z. Sienna Pan, Tanzil Rujedawa, Benjamin W. Lamb, Laura Succony, Shi Lam, Bincy M. Zacharia, Rebecca Lucey, Alexander J. P. Fulton, Dimana Kaludova, Anita Balakrishnan, Juliet A. Usher-Smith, Grant D. Stewart

## Abstract

**Objectives:** To identify and compare guidelines which make recommendations surveillance for the detection of recurrence for 16 common solid cancers after initial treatment with curative intent in asymptomatic patients.

**Design:** We conducted a systematic review, combining search results from two electronic databases (MEDLINE, EMBASE) and one guideline organisation website (NICE), as well as using expert consultation and manual searching. Screening and data extraction were carried out by multiple reviewers. We collected data from each guideline on the recommendations for surveillance and the use of risk-stratification. Findings were compared between cancer types and regions. Text mining was used to extract statements commenting on the evidence for surveillance.

**Results:** We identified 123 guidelines across 16 cancer types. Almost all guidelines (*n=*115, 93.5%) recommend routine surveillance for recurrent disease in asymptomatic patients after initial treatment. Around half (n=59, 51.3%) recommend indefinite or lifelong surveillance. The most common modality of surveillance was cross-sectional imaging. Risk-stratification of the frequency, length, and mode of surveillance was widespread, with most of the guidelines (*n=*92, 74.8%) recommending that surveillance be adapted based on assessment of patient risk. More than a third of the included guidelines (*n=*50, 39.0%) provided incomplete or vague recommendations about surveillance. For fourteen of the included cancers, we found statements in the guidelines indicating that there is no evidence that surveillance improves survival.

**Conclusions:** Although specific details of follow-up schedules vary, common challenges were identified across the 16 included cancer types. These include heterogeneity between recommendations for the same cancer type, vague or non-specific recommendation and a lack of cited evidence to support use of surveillance to improve outcomes for patients. Challenges in generating evidence in this area remain, however, increased availability to linked health records may provide a way forward for researchers.

**Registration:** Protocol published on PROSPERO in 2021 (ID: CRD42021289625)

## Introduction

Large numbers of people are diagnosed with cancer every year (18 million in 2020), and incidence is expected to continue rising in coming decades (28 million in 2024) [1]. In recent years, improved treatments have resulted in improved survival outcomes; in England improvements to treatment have contributed to the 7.8% increase in five-year survival rates between 2005 and 2016 [2]. Combined, these trends result in increasing numbers of people living with and beyond a cancer diagnosis; it is estimated that by 2030 this will include four million people in the UK [3].

Surgery is the first line of treatment for most solid non-metastatic cancers (either alone or in combination with adjuvant therapies). In England, 67% of treated cancers are managed surgically [4]. After initial treatment, patients transition into follow-up care, which may include surveillance to detect recurrent cancer before the emergence of symptoms or further spread of disease. Intensive surveillance, for example regular CT scans, is resource intensive for healthcare systems and places a high burden on patients. In a recent analysis of kidney cancer surveillance, using a multi-centre European cohort, it was estimated that 542 CT scans were required to detect one curable recurrence [5].

In this systematic review, we identified guidelines which include recommendations for surveillance to detect recurrence in asymptomatic individuals following surgical cancer treatment with curative intent. We included all 16 solid cancers in the list of 20 most common incident cancers in the UK [6]: breast, prostate, lung, bowel (colorectal), melanoma, kidney, head and neck, brain and central nervous system (referred to as brain hereafter), pancreas, bladder, uterus (endometrial), oesophagus, ovary, stomach (gastric), liver and thyroid. We compare identified guidelines to assess heterogeneity within and between cancer types, looking in detail at the types of surveillance recommended, the length of follow-up and the use of risk stratification. We also examine the evidence presented by the guidelines that surveillance improves outcomes for patients.

## Methods

We performed a systematic review following an *a priori* established study protocol (PROSPERO 2021 CRD42021289625).

Guidelines for inclusion were identified through a three-stage process. In the first stage, an electronic literature search of Medline, EMBASE and the National Institute for Health and Care Excellence (NICE) website search engine was performed (November 2021). Note that the NICE search engine captures publications from a range of guidelines bodies (including but not limited to NICE guidelines). We included literature published in 2010-2021, using a combination of subject headings incorporating “cancer/neoplasm” and “follow-up/surveillance”, limited to publications classified as guidelines. The full search strategy is provided in the supplementary materials.

We included guidelines that fulfilled the following criteria:

- Describe recommendations, strategies or information which can assist clinicians and patients to make decisions about routine surveillance (type, frequency, length) in asymptomatic adult patients after treatment with surgery with curative intent for one of the 16 included solid cancers.
- Produced by medical specialty associations, including relevant professional societies, public or private organisations, government agencies or healthcare providers at the state, national or international level.
- Freely available in print or electronic format in English.

Guidelines developed for specific populations (including groups with rare genetic conditions), less common cancer sub-types (<10% of incident cases of one of the listed solid cancers) or metastatic disease only were excluded.

One reviewer carried out the electronic database search (HH) and another carried out the National Institute for Health and Care Excellence (NICE) website search (BKS). Two reviewers independently screened each publication identified in the searches by title and abstract to exclude clearly irrelevant results (BKS, FK, HH). If a definite decision to exclude could not be made, two reviewers independently screened the full text (BKS, FK, HH). Disagreements were resolved by discussion with the third reviewer.

In the second stage, clinical subject experts were consulted and asked to identify additional guidelines currently in use within their speciality. We consulted 13 UK-based clinical experts (identified through clinical networks) and sent them each a questionnaire with information about the guidelines identified in the first stage relating to their speciality (or specialities). Additionally, manual checking of well-known guideline organisations (including the National Comprehensive Cancer Network (NCCN), European Society for Medical Oncology (ESMO), and American Society of Clinical Oncology (ASCO)) was carried out by one reviewer (HH). All guidelines identified in the second stage were also screened by two reviewers independently (HH, BKS) to assess eligibility for inclusion.

Finally, in the third stage, to ensure the final set of results provided an up-to-date overview of the available documents, checks for updates to each guideline were carried out in April 2023 by one reviewer (HH).

A standardised data extraction form was developed by the study team and was piloted for the identified guidelines for one cancer type (kidney). For each included guideline, one reviewer extracted the data into the form, and a second reviewer checked the results (HH, BKS, FK, SHR, CR, LS, SL, BMZ, BWL, RL, AJPF, DK, EW, ZSP, EG, SKS, AE, TR and AB). Information about the population covered by the guideline, details of recommendations for routine surveillance in asymptomatic patients following curative treatment (including modality and length) and the use of risk-stratified surveillance were recorded. During data extraction, reviewers also identified any recommendations for surveillance that were vague, incomplete, or non-specific. A narrative synthesis approach was used to identify similarities and differences between cancer types and regions.

To identify statements in the guidelines describing evidence that surveillance affects clinical outcomes, a text mining approach was used to identify sentences in the guideline documents containing words relating to evidence (including “evidence” and “data”) alongside words relating to follow-up or relevant outcomes (including “survival”, “mortality” and “follow-up”). Sentences containing words specific to the treatment or diagnosis of cancer (“adjuvant”, “chemotherapy” and “preoperative”) were removed. The resulting sets of sentences were then manually screened.

Patients and members of the public have not been involved in this research study.

## Results

After the removal of duplicates, the search of electronic databases and the NICE website identified 6385 publications, 6223 of which were excluded through title and abstract screening (*n=*162). A further 80 publications were identified through citation searching (*n=*13), consultation with experts (*n=*32) and manual checking of guideline organisations (*n=*35). These, 242 publications in total, were screened by full text, of which 121 were found to meet the review criteria. Five guidelines were replaced by an update from the same guideline body following checks in April 2023. Two identified guidelines cover two cancer types (gastric and oesophageal), so we subsequently refer to 123 included guidelines. A PRISMA flow chart (Fig. 1), a list of included studies (Table S1a) and a list of studies excluded in full text screening (Table S1b) are provided.

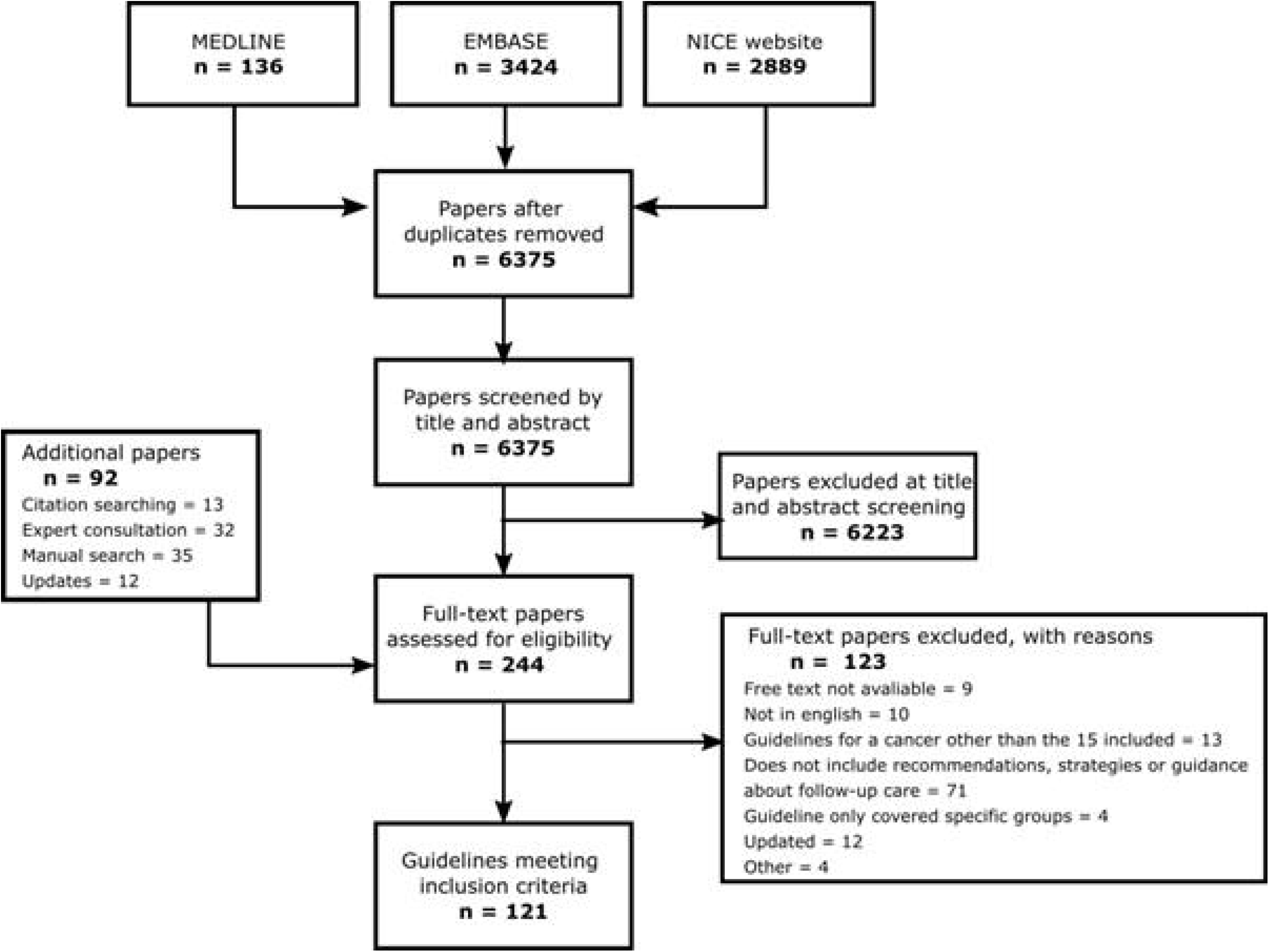

At least three guidelines were identified for each of the included cancer types (Table 1, Table S2, Table S3a). The largest number of guidelines were identified for colorectal cancer (*n*=18, 14.6%) and the fewest for brain cancer (*n*=3, 2.4%). The identified guidelines were produced by 51 medical speciality organisations covering: well-known cancer guideline organisations (including NCCN (*n*=16, 13.0%), ESMO (*n*=16, 13.0%) and ASCO (*n*=6, 4.9%)); speciality organisations for specific cancer types (including the European Association of Urology (EAU) (*n*=3, 2.4%) and American Thyroid Association (ATA) (*n*=1, 0.81%)); and from national organisations providing guidance for healthcare systems (including NICE (*n*=12, 9.8%) and Cancer Council Australia (CCA) (*n*=3, 2.4%)). The guidelines covered a variety of geographic regions (Table S3b); however, the vast majority gave recommendations for populations in Europe (*n*=59, 48.0%), including 20 (16.3%) from the UK, and North America (*n*=46, 37.4%), of which the majority (*n*=41, 33.3%) were from the USA. We identified a small number of guidelines developed for populations in Asia (*n*=7, 5.7%), Oceania (*n*=5, 4.1%), South America (*n*=4, 3.3%) and the Middle East (*n*=1, 0.8%). Two guidelines were identified that did not relate to a specific region. No guidelines for countries or regions in Africa were identified.

**Table 1:** Summary of surveillance recommendations by cancer type.

| Cancer Type | Number | Is surveillance to detect recurrence recommended? |  |  |  |  | Who is recommended cross-sectional imaging as part of routine surveillance? |  |  |  | Fixed end point for surveillance |
| --- | --- | --- | --- | --- | --- | --- | --- | --- | --- | --- | --- |
|  |  | Any (%) | Imaging (%) |  | Biomarkers (%) | Clinical App (%) | Everyone (%) | Some individuals (%) | Nobody (%) | Unclear (%) |  |
|  |  |  | Cross-sectional | Localised |  |  |  |  |  |  |  |
| Overall | 123 | 115 (93.5) | 89 (72.4) | 68 (55.3) | 55 (44.7) | 71 (57.7) | 34 (27.6) | 45 (36.6) | 32 (26) | 12 (9.8) | 25 (20.3) |
| Bladder | 10 | 10 (100) | 10 (100) | 10 (100) | 9 (90.0) | 2 (20.0) | 1 (10.0) | 9 (90.0) | - | - | 1 (10.0) |
| Brain (and CNS) | 3 | 3 (100) | 3 (100) | - | - | 2 (66.7) | 3 (100) | - | - | - | - |
| Breast | 9 | 9 (100) | 9 (100) | 3 (33.3) | - | 7 (77.8) | 9 (100) | - | - | - | 4 (44.4) |
| Colorectal | 18 | 18 (100) | 16 (88.9) | 16 (88.9) | 13 (72.2) | 11 (61.1) | 7 (38.9) | 7 (38.9) | 2 (11.1) | 2 (11.1) | 4 (22.2) |
| Endometrial | 4 | 4 (100) | 2 (50.0) | - | - | 4 (100) | - | 2 (50.0) | 2 (50.0) | - | 3 (75.0) |
| Gastric | 7 | 5 (71.4) | 4 (57.1) | 4 (57.1) | 2 (28.6) | 2 (28.6) | 1 (14.3) | 3 (42.9) | 3 (42.9) | - | 2 (28.6) |
| Head and Neck | 6 | 6 (100) | 2 (33.3) | 5 (83.3) | - | 4 (66.7) | - | 3 (50.0) | 3 (50) | - | - |
| Kidney | 7 | 7 (100) | 7 (100) | 5 (71.4) | 4 (57.1) | 3 (42.9) | 7 (100) | - | - | - | 1 (14.3) |
| Liver | 5 | 5 (100) | 4 (80.0) | - | 3 (60.0) | 1 (20.0) | 4 (80.0) | - | - | 1 (20.0) | 1 (20.0) |
| Lung | 6 | 6 (100) | 5 (83.3) | - | - | 5 (83.3) | - | - | 1 (16.7) | 5 (83.3) | - |
| Melanoma | 15 | 15 (100) | 14 (93.3) | 12 (80.0) | 6 (40) | 14 (93.3) | - | 13 (86.7) | 1 (6.7) | 1 (6.7) | 6 (40.0) |
| Oesophageal | 7 | 5 (71.4) | 4 (57.1) | 4 (57.1) | - | 3 (42.9) | 1 (14.3) | 3 (42.9) | 3 (42.9) | - | 2 (28.6) |
| Ovarian | 7 | 5 (71.4) | 1 (14.3) | 2 (28.6) | 2 (28.6) | 5 (71.4) | - | - | 6 (85.7) | 1 (14.3) | 1 (14.3) |
| Pancreas | 4 | 2 (50.0) | 2 (50.0) | - | 2 (50.0) | 2 (50.0) | 1 (25.0) | - | 2 (50.0) | 1 (25.0) | - |
| Prostate | 8 | 8 (100) | - | - | 8 (100) | 3 (37.5) | - | - | 8 (100) | - | - |
| Thyroid | 7 | 7 (100) | 6 (85.7) | 7 (100) | 6 (85.7) | 3 (42.9) | - | 5 (71.4) | 1 (14.3) | 1 (14.3) | - |

### Recommendation for surveillance

Of the guidelines assessed, almost all (*n*=115, 93.5%) recommend at least some form of routine surveillance for recurrent disease in asymptomatic patients after initial surgery (Table 1). Eight guidelines (covering four cancer types) recommend against routine surveillance for asymptomatic patients:

- Two European guidelines for ovarian cancer (published in 2011 and 2013). These pre-date the widespread use of the biomarker cancer antigen 125 (CA125) which is recommended as part of routine surveillance for the recurrence of ovarian cancer by ESMO guidelines from 2019.
- The two UK-based guidelines for oesophageal and gastric cancer. This contrasts with the guidelines developed for these two cancers by the pan-European ESMO as well as organisations in the USA, South America, Asia, and the Middle East.
- The two guidelines for pancreatic cancer developed by European guideline bodies. The other two guidelines identified for pancreatic cancer, which do recommend routine surveillance in asymptomatic patients, were developed by guideline bodies in the USA.

### Duration of surveillance

Of the 115 guidelines which recommend some form of surveillance, most (n=81, 70.4%) do not recommend a fixed end point (Table S2, Table S3a). Six explicitly recommend lifelong surveillance (including 3 of the 15 melanoma guidelines) and 53 recommend that surveillance should be indefinite for all patients (including 5 of the 6 lung cancer guidelines). There is no mention of the expected length of surveillance in 22 of the guidelines.

Of the remaining 34 guidelines, 25 recommend a fixed duration of surveillance for all patients. This includes three (75%) of the guidelines for endometrial cancer, four for breast cancer (44.4%) and six for melanoma (40%). Of these 25, most (*n*=19, 76.0%) recommend stopping surveillance for all patients by five years and the remainder recommend stopping by 10 years. No guidelines for brain, head and neck, lung, pancreas, prostate or thyroid cancers and only one guideline for bladder, kidney, liver and ovarian cancers give fixed durations for follow-up for all patients.

The nine remaining guidelines give a fixed duration for some patients and an indefinite (or lifelong) duration for others (using a risk-stratified approach). This includes seven of the guidelines for bladder cancer (70%), which advise adjusting the length of follow-up based on risk of recurrence. For example, the EAU guidelines, which recommend that those at lowest risk of recurrence should stop follow-up after five years while those at highest risk should have lifelong follow-up.

### Modalities of surveillance

The most common surveillance modality is cross-sectional imaging (Table 1, Table S4a). This is recommended for at least some patients by 89 (72.4%) of the guidelines. There are four cancers (bladder, brain, breast, and kidney) for which all the identified guidelines recommend cross-sectional imaging as part of routine surveillance. There are seven further cancers in which most guidelines recommend this type of imaging for at least some asymptomatic patients during routine surveillance: melanoma (93.3%), colorectal (88.9%), thyroid (85.7%), lung (83.3%), liver (80%), oesophageal (57.1%) and gastric (57.1%). None of the prostate cancer and only one of the guidelines for ovarian cancer recommend the use of this type of imaging (instead relying mainly on Prostate Specific Antigen (PSA), or a combination of gynaecological exams and CA125 testing respectively).

There are differences in recommendations for the use of cross-sectional imaging for surveillance between different regions (Tables S3b-d). For colorectal, endometrial, gastric, head and neck, liver, lung, oesophageal, ovarian, pancreatic, and thyroid cancers, the proportion of European guidelines recommending this type of imaging is lower than the proportion of North American guidelines making the same recommendation. For ovarian and pancreatic cancers, the only recommendations for cross-sectional imaging are made by guidelines from the USA (*n*=1, 14.3% and *n*=2, 50% respectively). For lung and thyroid cancers, guidelines from the UK are the only ones which do not make a recommendation for cross-sectional imaging. Overall, the proportion of the guidelines from North America recommending at least some cross-sectional imaging during follow-up is substantially higher (*n*=39, 86.7%) than European guidelines (*n*=35, 60.3%).

The most common modality of cross-sectional imaging is computed tomography (CT) scans, recommended for use in surveillance in more than half of the identified guidelines (*n*=67, 54.5%) (Table S4a). Almost all guidelines for bladder (*n*=9, 90.0%), colorectal (*n*=15, 83.3%), kidney (*n*=7, 100%), liver (*n*=4, 80.0%) and lung (*n*=5, 83.3%) recommend using CT scans in surveillance for at least some patients, whereas for breast and brain cancers, no guidelines recommend the use of CT scans (instead mammograms and MRI are recommended respectively). Five different cross-sectional imaging modalities are recommended across the 15 melanoma guidelines (CT scans (*n*=11), MRI (*n*=3), PET (*n*=4), multimodal (*n*=5) and X-ray (*n*=2)).

Localised imaging techniques are also widely recommended for surveillance (*n*=68, 55.3%) (Table 1, Table S4b). For bladder and thyroid cancer, all identified guidelines recommend localised imaging for at least some of the individuals undergoing routine surveillance (cystoscopy and ultrasound respectively). In contrast, none of the guidelines for brain, endometrial, liver, lung, pancreatic or prostate cancer recommend imaging of this type. These cancers rely on cross-sectional imaging (brain, liver, lung and pancreatic), biomarkers (pancreatic and prostate), physical exams (endometrial, lung, pancreatic and prostate) or symptom monitoring (brain, lung and pancreatic).

Ten cancer types have at least one guideline that recommends using a biomarker within routine surveillance (*n*=55, 44.7%) (Table 1, Table S4b). Biomarkers are recommended in most guidelines for six cancer types: bladder (*n*=9, 90%), colorectal (*n*=13, 72.2%), kidney (*n*=4, 57.1%), liver (*n*=3, 60%), prostate (*n*=8, 100%) and thyroid (*n*=6, 85.7%). Carcinoembryonic antigen (CEA) is recommended for use in surveillance of recurrence by 13 (72.2%) colorectal cancer guidelines, all prostate cancer guidelines (*n*=8) recommend the use of PSA and nine guidelines (90%) for bladder cancer recommend the use of urine cytology. For ovarian cancer, two guidelines (28.6%) recommend the use of CA125 in surveillance for recurrence, however, two other guidelines explicitly recommend against its use in this context, citing lack of evidence. Four guidelines for kidney cancer (57.1%) recommend the use of various biomarkers to monitor kidney function during routine surveillance, however, none recommend a biomarker that can detect recurrence.

A follow-up appointment with a clinician is explicitly recommended in at least one guideline for each cancer type (Table 1, Table S2). This includes most guidelines for brain (*n*=2, 66.7%), breast (*n*=7, 77.8%), colorectal (*n*=11, 61.1%), endometrial (*n*=4, 100%), head and neck (*n*=4, 66.7%), lung (*n*=5, 83.3%), melanoma (*n*=14, 93.3%) and ovarian (*n*=5, 71.4%) cancers. Physical exams are recommended by 62 guidelines, although the specifics vary between cancer types. All endometrial and five (71%) ovarian cancer guidelines recommend a gynaecological exam; 12 (80%) melanoma guidelines recommend a physical exam, with several (*n*=5) explicitly mentioning a skin exam or body mapping; 11 (61.1%) colorectal cancer guidelines recommend a physical exam, of which most (*n*=8, 72.70%) specify a rectal exam. History taking (including discussion of symptoms) is recommended by 45 guidelines, including most of the guidelines for lung (*n*=5, 83.3%), endometrial (*n*=3, 75.0%), breast (*n*=6, 66.7%) and brain (*n*=2, 66.7%) cancers. Only a small number of guidelines for gastric (*n*=2, 28.6%), bladder (*n*=2, 20%) and liver (*n*=1, 20%) cancers mention a clinical appointment.

Around 10% of guidelines (*n*=12, 9.8%) recommended a non-specific modality of surveillance. For example, recommendations that imaging should be part of routine surveillance without further details.

### Risk-stratification and surveillance

The use of risk-stratification during follow-up after surgery for cancer is widely recommended, with 92 of the included guidelines (74.8%) - and at least one guideline for each of the 16 cancers - recommending that different groups of patients receive different routine surveillance (Table 2, Table S3a). Notably, all guidelines for four cancer types (bladder, head and neck, kidney, and thyroid) recommend the use of risk-stratification.

**Table 2:** Summary of risk-stratification recommendations by cancer type.

| Cancer Type | Number | Is risk-stratification recommended? |  |  |  |  |  |  |
| --- | --- | --- | --- | --- | --- | --- | --- | --- |
|  |  | Any (%) | What is adapted? |  |  |  | How? |  |
|  |  |  | Eligibility (%) | Type (%) | Frequency (%) | Length (%) | Clinical risk factors (%) | Non-clinical risk factors (%) |
| **Overall** | 123 | 92 (74.8) | 6 (4.9) | 63 (51.2) | 62 (50.4) | 32 (26) | 83 (67.5) | 42 (34.1) |
| **Bladder** | 10 | 10 (100) | - | 9 (90.0) | 7 (70.0) | 7 (70.0) | 10 (100) | 2 (20.0) |
| **Brain and CNS** | 3 | 2 (66.7) | - | - | 2 (66.7) | 1 (33.3) | 2 (66.7) | 2 (66.7) |
| **Breast** | 9 | 5 (55.6) | - | 2 (22.2) | 3 (33.3) | 1 (11.1) | 4 (44.4) | 5 (55.6) |
| **Colorectal** | 18 | 15 (83.3) | 2 (11.1) | 11 (61.1) | 7 (38.9) | 7 (38.9) | 12 (66.7) | 8 (44.4) |
| **Endometrial** | 4 | 3 (75.0) | - | 2 (50.0) | 2 (50.0) | - | 3 (75.0) | - |
| **Gastric** | 7 | 5 (71.4) | 1 (14.3) | 3 (42.9) | 3 (42.9) | 2 (28.6) | 5 (71.4) | 3 (42.9) |
| **Head and Neck** | 6 | 6 (100) | - | 3 (50.0) | 1 (16.7) | 1 (16.7) | 4 (66.7) | 4 (66.7) |
| **Kidney** | 7 | 7 (100) | - | 5 (71.4) | 7 (100) | 3 (42.9) | 6 (85.7) | 4 (57.1) |
| **Liver** | 5 | 4 (80.0) | - | - | 3 (60.0) | - | 4 (80.0) | - |
| **Lung** | 6 | 4 (66.7) | - | 1 (16.7) | 3 (50.0) | - | 3 (50.0) | 2 (33.3) |
| **Melanoma** | 15 | 14 (93.3) | - | 14 (93.3) | 12 (80.0) | 3 (20.0) | 14 (93.3) | 6 (40.0) |
| **Oesophageal** | 7 | 4 (57.1) | 1 (14.3) | 4 (57.1) | 3 (42.9) | 3 (42.9) | 4 (57.1) | 1 (14.3) |
| **Ovarian** | 7 | 2 (28.6) | - | 1 (14.3) | 2 (28.6) | 1 (14.3) | 2 (28.6) | - |
| **Pancreas** | 4 | 1 (25.0) | - | 1 (25.0) | 1 (25.0) | - | 1 (25.0) | 1 (25.0) |
| **Prostate** | 8 | 3 (37.5) | - | 1 (12.5) | 1 (12.5) | 1 (12.5) | 2 (25.0) | 2 (25.0) |
| **Thyroid** | 7 | 7 (100) | 2 (28.6) | 6 (85.7) | 5 (71.4) | 2 (28.6) | 7 (100) | 2 (28.6) |

Many guidelines (*n*=63, 51.2%) recommend using stratification to determine the modality of surveillance. At least one guideline for 14 cancers (none for brain or liver cancer) stratifies in this way. Of the 89 guidelines that recommend the of use cross-sectional imaging, around half (*n*=46, 51.7%) explicitly limit usage to higher-risk individuals. This varies by cancer type: most of the guidelines for bladder (*n*=9, 90.0%), melanoma (*n*=13, 86.7%) and thyroid (*n*=6, 85.7%) recommend targeting the use of cross-sectional imaging, but three cancer types (brain, breast, and kidney) recommended some cross-sectional imaging for all patients.

At least one guideline for each cancer type recommends scheduling more frequent follow-up for patients at higher risk, including all seven kidney cancer guidelines and 12 (80.0%) of the melanoma guidelines. However, in 17 (out of 62) the details of the risk-adjusted surveillance schedules are incomplete. For example, three guidelines for gastric cancer (42.9%) recommend that higher risk patients should have more frequent follow-up, but the recommended frequency is not stated.

Most of the guidelines recommending risk-stratification (*n*=83, 90.2%), and at least one guideline for each cancer type, use clinical risk factors (i.e. characteristics of the cancer being treated) to assess risk of recurrence and determine appropriate surveillance. Stage is the most widely mentioned clinical risk factor (*n*=63, 68.5%), used by guidelines for 14 cancer types, including almost all the guidelines for bladder (*n*=8, 80.0%), kidney (*n*=6, 85.7%), and thyroid cancers (*n*=6, 85.7%). Non-clinical risk factors, including demographic and lifestyle risk factors, are used in nearly half of the guidelines recommending risk stratification (*n*=42, 45.7%) and across 13 cancer types. The use of risk factors such as age, eligibility for further treatment or frailty, in several guidelines (*n*=28, 30.4%) indicates consideration of competing risks (such as death from other causes) when assessing patients. Some guidelines use risk factors for the development of the cancer type they cover (for example, smoking is mentioned by three guidelines for head and neck cancer) to estimate risk of recurrent disease.

Nearly a third of guidelines (*n*=39, 31.7%) give an incomplete description of the recommended method of risk-stratification – including at least one guideline for eleven of the included cancer types. This is seen in seven of the melanoma guidelines (46.7%); clinicians are advised to classify patients as high-risk if they have “high risk pathologic features” or “risk factors for melanoma development”.

### The Evidence-basis for surveillance

Overall, 78 guidelines (63.4%) presented some evidence or assessed the evidence level for at least one aspect of their follow-up recommendation (Table S2, Table S3a); the remaining guidelines made recommendations based only on expert opinion (*n*=31, 25.2%) or did not state how the recommendations were reached (n=14, 11.4%).

Using a text mining approach, we identified 45 statements in guidelines for 14 cancer types which discuss the evidence that surveillance improves long-term outcomes for patients (Table S5). No statements of this type were identified in any of the guidelines for endometrial, liver, or thyroid cancers.

“…*older meta-analyses have reported an improvement in overall survival in intensively surveilled populations*.*” (European Society of Gastrointestinal Endoscopy and European Society of Digestive Oncology, Colorectal Cancer Guideline, 2019)*

“*However, at this time, no data from prospective trials demonstrating the potential benefit of early detection of recurrent disease and its impact on OS are available*.*” (European Association of Urology, Bladder Cancer Guideline, 2021)*

“*There is currently no data demonstrating improvements in survival from routine surveillance*.*” (Cancer Care Ontario, Lung Cancer, 2014)*

“*Although there is no research showing that periodic imaging lengthens the OS, many countries’ guidelines recommended periodic imaging following curative resection of melanoma” (Japanese Dermatological Association, Melanoma Cancer Guideline, 2020)*

*Box 1: statements on surveillance and survival (selected extracts)*

We identified statements in two colorectal cancer guidelines asserting that intensive surveillance following surgery can improve overall survival (Box 1). Conversely, we identified sentences in guidelines for two cancers (pancreatic, and head and neck) stating that there is no evidence for surveillance, in guidelines for three cancers (bladder, brain and pancreatic) that there is no benefit to surveillance and in guidelines for nine cancers (bladder, breast, colorectal, gastric, lung, melanoma, oesophagus, ovarian and prostate) that there is no evidence that surveillance improves outcomes (in most cases survival is specified). One guideline for breast cancer also said that there is no palliative benefit to the use of surveillance to detect recurrences earlier. Two guidelines (for breast and lung cancer) emphasised that asymptomatic detection of recurrence is unnecessary if patients have access to healthcare if symptoms emerge.

“*Intensive follow-up can detect recurrences earlier, thus surgical resection for curative intent is possible. However, this is not associated with improved survival*.*” (Cancer Council Australia, Colorectal Cancer Guideline, 2018)*

“*Although the hypothesis suggesting that regular monitoring reveals early detection of metastasis may be well founded, no randomised studies have demonstrated that early detection of metastases improves the overall survival*.*” (Swiss Guideline, Melanoma Guideline, 2016)*

“*A regular follow-up may allow investigation and treatment of symptoms, psychological support and early detection of recurrence, though there is no evidence that it improves survival outcomes” (European Society for Medical Oncology, Gastric Cancer Guideline, 2016)*

*Box 2: statements on surveillance and early detection (selected extracts)*

Several guidelines discuss the link between surveillance, earlier diagnosis of recurrence and survival outcomes (Box 2). Four guidelines for colorectal cancer state that surveillance improves early detection (or increases the number of treatable recurrences), while guidelines for melanoma and gastric cancers state that this improvement is expected. Guidelines for breast and lung cancer referred to evidence that early detection and early treatment of recurrence are associated with improved survival. However, guidelines for seven cancers (brain, colorectal, gastric, melanoma, oesophagus, ovarian and prostate) said that there is not currently any evidence that earlier detection of recurrence improves survival, and guidelines for three cancer types (kidney, ovarian and pancreatic) similarly claim that the earlier treatment of advanced disease does not improve survival (Box 3). One guideline for ovarian cancer went further, citing evidence that treating recurrences early is associated with a decrease in quality of life.

“*It is important to underscore the goal of surveillance in early-stage breast cancer, which is to detect early locoregional or contralateral recurrence, as early detection of breast cancer recurrence is correlated with improved survival*.*” (American College of Radiology, Breast Cancer Guideline, 2019)*

“*Nevertheless, there are no trials indicating that the earlier detection of recurrence and subsequent change in management improves outcomes*. “ *(European Society for Medical Oncology, Prostate Cancer Guideline, 2020)*

“*The data suggest that treating recurrences early (based on detectable CA-125 levels in patients who are asymptomatic) is not associated with an increase in survival and is associated with a decrease in QOL” (National Comprehensive Cancer Network, Ovarian Cancer Guideline, 2023)*

“…*there is no evidence to show whether early detection leads to improved overall survival*.*” (National Institute for Health and Care Excellence, Gastric Cancer Guideline, 2018)*

*Box 3: statements on early detection (or treatment) of recurrence and survival (selected extracts)*

Additionally, statements indicating a lack of evidence about the specific protocols or schedules for surveillance being recommended were identified in at least one guideline for 15 of the included cancer types (Box 4, Table S5). No statements on this topic asserted that there was any evidence favouring one surveillance schedule over another.

“…*there are no differences in overall survival with any of the different follow-up schemes… if the patient is guaranteed access to healthcare services in the presence of a symptom or sign of alarm*.*” (European Society for Medical Oncology, Breast Cancer Guideline, 2019)*

“*Local follow-up protocols are based more on historical practice than evidence and are often disease-rather than patient-centred*.*” (NICE, Head and Neck Cancer Guideline, 2016)*

““*However, the ideal monitoring intervals and methods require further research*.*” (Korean Liver Cancer Association–National Cancer Center Korea, Liver Cancer Guideline, 2019)”*

“*There is no high-quality evidence to support one surveillance schedule for follow-up care of melanoma survivors, which results in great variability in guideline recommendations from different organizations*.*” (Cancer Care Ontario, Melanoma Cancer Guideline, 2015)*

*Box 4: statements on evidence basis for specific surveillance protocols or schedules (selected extracts)*

## Discussion

### Summary

In this first multi-cancer review we identified 123 guidelines for the surveillance of asymptomatic patients following curative surgery across 16 solid cancer types, finding that almost all guidelines recommend surveillance for recurrence in asymptomatic individuals (*n*=115, 93.5%). Where recommendations for surveillance were found, we noted substantial differences between guidelines both within and between cancer types, widespread inclusion of vague or non-specific recommendations and an almost complete absence of cited evidence that surveillance improves long-term patient cancer outcomes.

### Challenges Identified

The heterogeneity observed between guidelines for the same cancer type was notable. Differences are observed in modality (five cross-sectional imaging modalities are recommended across the melanoma guidelines) and scheduling (the recommended length of surveillance for oesophageal cancer ranges from two years to indefinite) of surveillance, as well as in the use of risk-stratification. There are notable exceptions to this such as: the use of PSA and cystoscopy are recommended in all the guidelines for prostate and bladder cancers respectively. Some of the heterogeneity can be explained by changing clinical practise over time (such as the more widespread use of CA125 in recent ovarian cancer guidelines) or geographical variations in clinical practice. Across several cancer types cross-sectional imaging is less widely recommended in Europe (particularly UK guidelines) compared to North America. However, there are also differences in surveillance recommendations between guidelines in the same region for the same cancer type; there are four guidelines from the USA for prostate cancer, two recommend more surveillance for those at higher risk of recurrence while the other two recommend the same surveillance for everyone. Variation in the recommendations for follow-up care have been previously identified in reviews of guidelines conducted for prostate cancer [7] and melanoma [8], in 2009 and 2015 respectively, corroborating our findings and suggesting this is an ongoing challenge.

We found several areas in which recommendations are frequently vague and non-specific. Many guidelines (*n*=53) recommend indefinite surveillance and others (*n*=22) do not mention the expected length of the recommended surveillance. In both cases, decision-making about ending surveillance is left to individual clinicians. Patients who are not eligible for further treatment (for example, because they are too frail) are unlikely to see any benefit from continued surveillance for the early detection of recurrence. Although some guidelines (*n*=28, 22.8%) do recommend considering the age, frailty, or eligibility for further treatment of patients as part of their recommendation, we did not find recommendations encouraging periodic reassessment to ensure that surveillance continues to be appropriate. We also found that many guidelines recommending risk-stratification did not clearly define the risk factors they suggest using and others gave only vague details of how surveillance schedules should differ between high and low risk groups. This allows for variability in their interpretation by clinicians, which is likely to lead to variability in care. Lack of clear definitions of risk factors was also seen in a previous review of follow-up guidelines for melanoma [8].

Variation in the recommendations and a lack of specific details are likely connected to the limited evidence that surveillance improves patient outcomes, mentioned in guidelines for almost all cancer types. The reported lack of evidence is particularly concerning given that the use of surveillance to detect recurrence is so widespread. Many guidelines recommend lifelong or indefinite surveillance which presents a significant burden for both patients and healthcare systems. We highlight the frequent use of cross-sectional imaging, which typically involves exposure to radiation and utilises a valuable resource in terms of equipment and trained healthcare professionals. The use of risk-stratification, seen across all included cancer types, is a reasonable method to distribute resources among patients according to need, ideally improving rates of detection in high-risk patients and reducing burden for low-risk patients. However, we did not find any statements in guidelines discussing any evidence of the effect of a risk-stratified schedule on patient outcomes. Previous reviews of colorectal cancer [9] and melanoma [8] follow-up guidelines found a lack of evidence for the delivery of effective care and a need for research examining the benefits and costs for different follow-up strategies respectively.

### Strengths and weaknesses

This is the first review of follow-up guidelines for multiple cancer types, providing a comprehensive overview of this area of cancer care with the use of a rigorous search strategy (combining electronic databases, grey literature, consultation with subject experts and manual searching) to ensure good coverage from a range of guideline bodies. However, the decision to limit this large review to English language guidelines has led to an overrepresentation of guidelines from English speaking countries (including the UK, USA and Canada) and from international guideline bodies which publish in English (such as ESMO). It is our view, that this approach was sufficient to provide an overview of this topic – but results should be interpreted taking this into consideration.

The use of a text mining approach to identify statements about the evidence for surveillance in this review provided a pragmatic option to systematically identify relevant sentences across many lengthy documents. While data extraction typically focused on the subsection of the documents discussing follow-up, this allowed for the inclusion of text from throughout the guidelines. However, relevant statements may have been missed due to the formatting of the guideline documents or if they used language not captured by the search terms.

Researchers who are interested in exploring this topic further – for example, investigating in more detail the specific schedules of follow-up recommended or carrying out more detailed assessments of the evidence provided in the guidelines - may find the full list of included guidelines (including date, guideline body, URL) and the detailed extraction summary table (including details of the population covered by the guideline, surveillance modalities and risk stratification methods) provided in the supplementary materials (Tables S1a and S2) useful resources.

### Future Research

This review identified many guidelines which include recommendations on the use of surveillance to detect recurrent disease in asymptomatic patients following surgery for a solid cancer. We have identified areas in which recommendations for surveillance are often vague and non-specific, and shown that details of the recommendations vary between guidelines, including between guidelines for the same cancer type. While we recommend that guideline bodies consider these findings when updating their recommendations (with a focus on providing clear, specific advice that will help limit variation in care) we acknowledge that this is challenging while evidence in this area is lacking.

Although almost all included guidelines recommend lengthy surveillance, many of the guidelines acknowledge that there is little evidence that this improves long-term outcomes (including survival) for patients. Some studies, both observational [10] and trial based [11] (for melanoma and colorectal cancer respectively) have had success in generating evidence around optimal surveillance schedules; an ongoing UK based multi-centre trial [12] aims to produce evidence about the effect of follow-up on survival in patients after surgery for gastric cancer. Nevertheless, this area presents many challenges for evidence generation. The long timeframes required and potential difficulties in patient recruitment, make trials of alternative surveillance schedules prohibitively expensive and complex. However, the increasing availability of national datasets with many years of linked health records may present alternative options for researchers looking to model the performance and costs of surveillance recommendations, to investigate the association between surveillance and survival, and the opportunity to design optimised surveillance strategies.

What is already known on this topic?

- Surveillance of asymptomatic patients for recurrence is common after curative surgery for cancer.
- The goal of surveillance is to improve early detection rates of recurrent disease and hence improve survival.
- For many cancers, there are multiple guidelines which make recommendations about follow-up care in cancer patients.

What this study adds?

- Almost all guidelines for the 16 included solid cancers recommend surveillance after curative surgery, in many cases, this has an indefinite or lifelong duration.
- Recommendations are often vague or non-specific, which may lead to variation in the care delivered to patients.
- There is very little evidence presented in the guidelines that current recommendations for surveillance improve patient outcomes.

**Summary box**

## Supporting information

Search Strategy

## Data Availability

The authors will provide additional data collected during this study upon request, please email the corresponding author.

## Acknowledgements

We would like to thank Isla Kuhn for her support in developing the search strategy for this review. We also would like to thank the following subject experts for their guidance on the available guidelines for follow-up care in their respective fields: Benjamin Lamb, Anita Balakrishnan, Grant Stewart, Isla Kuhn, William Turner, Richard Mair, Daniel Leff, Justin Davies, Felipe Correia Martins, Sheraz Markar, Richard Shaw, Robert Rintoul, Amit Roshan and Sheila Fraser.

## Ethics approval

No ethical approval was required for this study.

## Transparency declaration

The lead author (HH) affirms that the manuscript is an honest, accurate, and transparent account of the study being reported; that no important aspects of the study have been omitted; and that any discrepancies from the study protocol have been explained.

## Credit Statement

Protocol development: BKS, HH, GDS, JAUS

Searching: HH, BKS

Screening: BKS, FK, HH

Data Extraction: HH, SHR, AE, SKS, EG, ZSP, EW, DK, AJPF, RL, BMZ, SL, LS, CR, GS, BKS, AB, BWL, FK

Analysis: HH

Writing the manuscript: HH, JAUS, GDS

Reviewing the manuscript: HH, JAUS, GDS, SHR, GS

Administrative: CB

