## Supplementary material for "Recommendations on surveillance for recurrence in asymptomatic patients following surgery in 16 common cancer types: a systematic review": Search Strategy

**Search Strategy (Embase, Medline and NICE)**

**Embase** <1974 to 2021 November 04> - search run on 05/11/2021

1	exp solid malignant neoplasm/	1791871

2	(cancer* or neoplasm* or carcinom* or tumo?r or melanom*).mp. [mp=title, abstract, heading word, drug trade name, original title, device manufacturer, drug manufacturer, device trade name, keyword heading word, floating subheading word, candidate term word]	5622856

3	exp cancer prognosis/ or exp monitoring/ or exp follow up/ or exp risk assessment/ or exp cancer risk/	3424724

4	(follow?up or suv?llance or manag* or stratif* or prognos*).mp. [mp=title, abstract, heading word, drug trade name, original title, device manufacturer, drug manufacturer, device trade name, keyword heading word, floating subheading word, candidate term word]	4406867

5	1 or 2	5695146

6	3 or 4	6815683

7	5 and 6	1708321

8	exp practice guideline/	618101

9	7 and 8	59488

10	exp clinical trial/ or cross-over studies/ or double-blind method/ or random allocation/ or randomized controlled trials as topic/ or single-blind method/	1879447

11	epidemiology/ or exp case control study/ or cohort analysis/ or case study/ or longitudinal study/ or retrospective study/ or prospective study/ or observational study/ or correlation study/ or cross-sectional study/	3250356

12	10 or 11	4763603

13	9 not 12	39133

14	guideline.ti,ab.	104343

15	13 and 14	3998

16	limit 15 to yr="2010 -Current"	3424

Ovid **MEDLINE**(R) and Epub Ahead of Print, In-Process, In-Data-Review & Other Non-Indexed Citations, Daily and Versions(R) <1946 to November 04, 2021> - search run on 05/11/2021

1	exp Neoplasms/	3565965

2	(cancer* or neoplasm* or carcinom* or tumo?r or melanom*).mp. [mp=title, abstract, original title, name of substance word, subject heading word, floating sub-heading word, keyword heading word, organism supplementary concept word, protocol supplementary concept word, rare disease supplementary concept word, unique identifier, synonyms]	4278744

3	exp risk assessment/ or exp cancer risk/ or exp Prognosis/	1975935

4	(follow?up or suv?llance or manag* or stratif* or prognos*).mp. [mp=title, abstract, original title, name of substance word, subject heading word, floating sub-heading word, keyword heading word, organism supplementary concept word, protocol supplementary concept word, rare disease supplementary concept word, unique identifier, synonyms]	2720291

5	1 or 2	4699608

6	3 or 4	3866412

7	5 and 6	1081557

8	limit 7 to guideline	922

9	limit 8 to yr="2010 -Current"	136

**NICE website search** (https://www.nice.org.uk/) carried out on 04/11/2021

“cancer” OR “carcinoma” OR “neoplasm” OR “tumour” OR “melanoma” OR “lymphoma” limited to results tagged as Guidance (2889 results).
